# Adverse Events Among People Living with HIV Receiving Dolutegravir-Based Regimens: Findings from a Six-Month Active Pharmacovigilance Study in Uganda

**DOI:** 10.1101/2025.08.15.25333743

**Authors:** Collins Ankunda, Sharon Namasambi, Curthbert Agolor, Susan Nakubulwa, Yvonne Karamagi, Ronald Mulebeke, Ivan Kasamba, John Lukoma, Goefrey Kalungi, Jude Emunyu, Allan Serwanga, Sheila Ampaire, Diana Nakitto Kesi, Victoria Nambasa, Barbara Mukasa, Helen Byomire Ndagije

## Abstract

**Background:** Dolutegravir (DTG)–based antiretroviral therapy (ART) is recommended as first⍰line treatment for people living with HIV (PLHIV) in Uganda. However, data on its safety profile in routine care remain limited. This study actively monitored adverse events (AEs) among PLHIV receiving DTG ⍰based regimens, assessing factors associated with their occurrence.

**Methods:** We conducted a prospective cohort study nested within a parent cohort from July 2021 to September 2022 a cross three Ugandan sites (urban, peri ⍰urban, and rural). Adults on DTG/lamivudine/tenofovir (TLD), either transitioning from prior ART regimens or initiated on TLD as first⍰line therapy, were followed for six months. AEs were systematically captured using structured interviews and classified by the Medical Dictionary for Regulatory Activities (MedDRA) System Organ Class. Univariable and multivariable logistic regression analyses identified predictors of AEs.

**Results:** Among 607 participants, 50 (8.0%) reported adverse events (AEs). Neuropsychiatric disorders were most common (2.5%), followed by cardiovascular (1.8%) and reproductive system disorders (1.5%). Other AEs, including gastrointestinal, eye, and skin disorders, were rare (<1%). In multivariable analysis, participants at site □3 and being single had significantly lower odds of AEs (aOR □0.04, 95%□CI□0.00–0.29, *p*□=□0.002), (a OR□0.45, 95%□CI□0.22–0.92, *p*□=□0.028). Prior exposure to Zidovudine-based regimens was associate d with increased odds of AEs (aOR □3.33, 95%□CI□1.43–7.75, *p*□=□0.005).

**Conclusion:** DTG⍰based ART regimens demonstrated a favourable safety profile, with AEs occurring infrequently and predominantly in neuropsychiatric, cardiovascular, and reproductive categories. Study site, Marital status and prior Zidovudine exposure were associated with AE risk, emphasizing the need for tailored monitoring, strengthened awareness in AE reporting and standardized pharm acovigilance across facilities.

## Background

Dolutegravir (DTG), an integrase strand transfer inhibitor (INSTI), has rapidly become a cornerstone of antiretroviral therapy (ART) for people living with HIV (PLHIV) globally due to its potent antiviral efficacy, high genetic barrier to resistance, and favorable safety profile [1,2]. Since its introduction, DTG-based regimens, commonly combined with tenofovir disoproxil fumarate(TDF) and lamivudine (TLD), have been widely adopted as first-line treatment in m any low- and middle-income countries (LMICs), including Uganda[3–5].

Despite the recognized benefits of DTG, concerns have emerged regarding its safety and tolerability in real-world settings, particularly in populations with diverse comorbidities and demographic characteristics. Clinical trials have reported relatively low rates of adverse events (AEs) associated with DTG; however, post-marketing pharmacovigilance and cohort studies have revealed cases of neuropsychiatric symptoms, weight gain, metabolic changes, and other unexpected toxicities[6–8]. Such adverse events can compromise adherence, quality of life, and long-term treatment success, emphasising the need for systematic and active safety monitoring[9,10,6].

Active pharmacovigilance, involving prospective data collection and close clinical follow-up, is critical to accurately identify, characterize, and quantify AEs associated with DTG regimens in routine care[11]. This is especially important in low and middle-incomecountries (LMICs) where the burden of HIV is highest, healthcare infrastructure may limit passive reporting, and patients often face complex social and clinical challenges[12]. Moreover, ART history and prior exposure to different regimens may influence AE risk, still, data remain limited on how these factors modify safety outcomes in populations transitioning to or initiating DTG[6].

In Uganda, national HIV treatment guidelines have incorporated DTG-based regimens as the preferred therapy since 2018, yet comprehensive data on the safety profile of DTG in Ugandan PLHIV remain sparse[5]. Understanding the real ⍰world safety profile of DTG is crucial to guide clinical decisions, optimize patient management, and inform national HIV programs on potential risks and mitigation strategies. This study, nested within a larger cohort, actively monitored six ⍰month adverse events among Ugandan PLHIV on TLD, comparing new initiators with those transitioning from other regimens. It also contributes to global pharmacovigilance efforts and supports WHO recommendations for ART regimens, while providing critical insights into adverse event incidence, types, and associated factors, thereby facilitating evidence ⍰based improvements in HIV care and treatment outcomes.

## Methods and materials

### Study Design and Duration

This study was nested within a parent prospective cohort[13] conducted from July 2021 to September 2022 to monitor the occurrence of adverse events among people living with HIV receiving tenofovir disoproxil fumarate/lamivudine/dolutegravir (TLD). Each participant was followed for six months. Participants were classified into two groups based on their ART history: the exposed group, comprising individuals on TLD who had previously received other ART regimens (zidovudine/lamivudine, tenofovir disoproxil fumarate/lamivudine, or abacavir/lamivudine plus a Non-Nucleoside Reverse Transcriptase Inhibitors (NNRTI), and the non ⍰exposed group, comprising ART ⍰naïve individuals initiating TLD as their first regimen.

### Study Population

The study included consenting adult males and females (18years+), on TLD with a fully suppressed viral load (target not detected) in the past six months and no history of diabetes. We excluded patients with established hyperglycemia or diabetes, detectable HIV viremia, unclear or untraceable ART regimen history, and those who were pregnant.

### Study Sites and Sampling

The study was carried out at three chosen locations: Mildmay Uganda Hospital (Site □1), Luwero Hospital (Site □2), and Nyimbwa Health Centre □IV (Site□3). These sites were carefully selected for their capacity to provide a representative sample of both rural and urban populations. Electronic lists from the Clinic Master and Electronic Medical Records (EMR) systems were obtained, identifying 9,919 patients on TLD across the three sites. Sampling proportionate to size was used to ensure sufficient representation across urban, peri-urban, and rural settings, with each site contributing participants according to its catchment population, as shown in Table □1.

### Data Collection, Management, and Analysis

Data were collected using clinician ⍰administered questionnaires at baseline and during two follow ⍰up visits, each three months apart. At every visit, participants were actively asked whether they had experienced any adverse events. Additional information was abstracted from ART registers, patient cards, and electronic medical record systems, then entered into the Open Data Kit (ODK) platform. Data were electronically cleaned, with paper records maintained as backups, and analyzed using Stata version 16. Sociodemographic and clinical characteristics were summarized and categorized, while types of AEs were classified according to the Medical Dictionary for Regulatory Activities (MedDRA) System Organ Class[14].

### Statistical Analysis Plan

Data were analyzed using Stata version □16. Descriptive statistics summarized participant characteristics as frequencies and percentages. Adverse events were summarized by MedDRA System Organ Class, with frequencies, proportions, 95% confidence intervals, and specific events reported. Univariable logistic regression assessed associations between potential predictors and the binary outcome (adverse event yes/no). The final analysis included 607 participants after excluding 21 with missing outcome data. Variables with p<0.2 in univariable analyses and those deemed clinically relevant were entered into a multivariable logistic regression model to obtain adjusted odds ratios with 95% confidence intervals. Model fit was assessed using likelihood ratio tests and pseudo R^2^, with significance at p<0.05.

### Ethical consideration

Ethical approval was obtained from the Mildmay Uganda Research Ethics Committee (# REC REF 0812-2020) and the Uganda National Council for Science and Technology (HS1273ES). Participants were treated with confidentiality, and written informed consent was obtained prior to enrollment in the study. Administrative clearance from the respective study sites was also obtained prior to implementation.

## Results

### Description of study participants

Majority of the participants were; female (n=419, 67%), aged 40 years and above (n= 327, 52%), of normal BMI (n= 347, 56%), previously exposed TDF NRTI backbone ART regimen (n=407, 65%), ART experienced for five years or more (n=404, 64%), and euglycemic at baseline (n=605, 96%) as shown in Table 2 below.

**Table 1:** Distribution of study participants per site.

| Study Site | Patients on TLD per Selected Site | Percentage contribution to the study sample size | Sample Size |
| --- | --- | --- | --- |
| Mildmay Uganda Hospital (MUgH) | 6,136 | (62%) | 390 |
| Luwero Hospital | 2,980 | (30%) | 188 |
| Nyimbwa HCIV | 802 | (8%) | 50 |
| **Total** | **9,919** | **100%** | **628** |

**Table 2:**
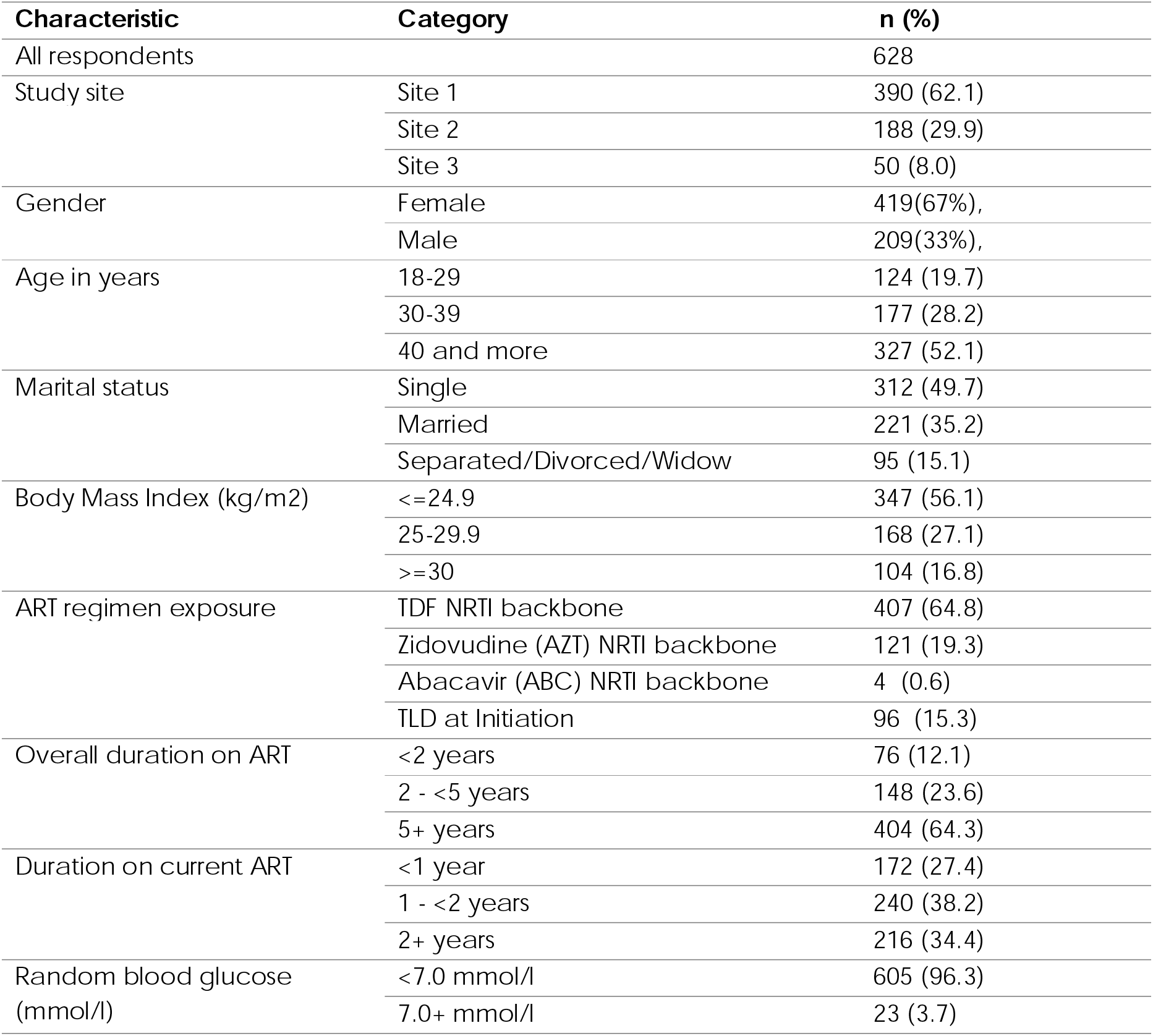
Baseline characteristics of study participants.

### Reported Adverse Events among PLHIV by MedDRA System Organ Classification

Among the 607 participants, adverse events were reported by 50 individuals (8.0%, 95% CI 6.1–10.4). The most frequently reported events were neuropsychiatric disorders (2.5%, 95% CI 1.5–4.1), predominant headaches (1.0%), peripheral neuropathy (0.7%), insomnia (0.3%), and forgetfulness (0.3%). Cardiac disorders followed (1.8%, 95% CI 1.0–3.2), mainly hypertension (1.6%). Reproductive system and breast disorders were also notable (1.5%, 95% CI 0.8–2.8), primarily erectile dysfunction (1.3%). Other events such as gastrointestinal (0.5%), eye disorders (0.7%), and skin-related symptoms (0.5%) were uncommon. Overall, the majority of participants (91.8%, 95% CI 89.3–93.7) reported no adverse events, indicating that most adverse effects were infrequent and concentrated in a few specific categories as shown in table 3 below.

**Table 3:**
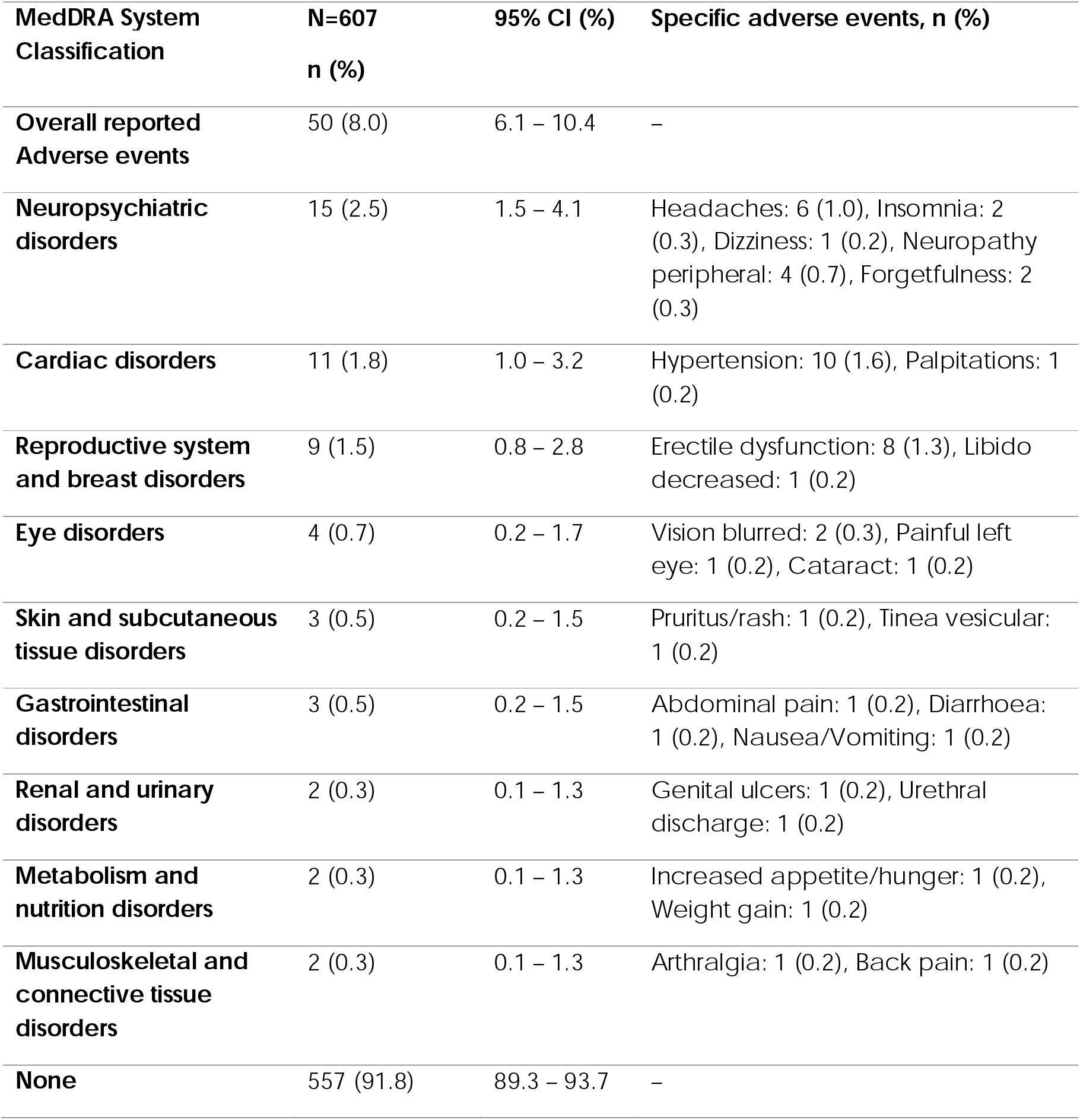
Distribution of Reported Adverse Events in PLHIV According to MedDRA Classification.

### Bivariate and Multivariate Analysis of Factors Associated with Adverse Events

Table 4 presents the univariable analysis, showing that participants at study site □3 had significantly lower odds of the outcome compared with those at site □1 (OR□0.04, 95%□CI□0.01–0.32, p□=□0.002). Being single was also associated with reduced odds (OR□0.51, 95%□CI□0.27–0.96, p□=□0.036), while having a BMI □≥□30□kg / m^2^ showed higher odds (OR□2.27, 95%□CI□1.09–4.72, p□=□0.028). Exposure to an AZT-based NRTI regimen was strongly associated with increased odds (OR □3.23, 95%□CI□1.72–6.08, p□<□0.001). In addition, a duration on current ART of ≥2□years was linked with higher odds (OR □2.24, 95%□CI□1.01–4.95, p□=□0.047). In multivariable analysis, site □3 remained signific ant with markedly reduced odds (aOR □0.04, 95%□CI□0.00–0.29, p□=□0.002). Being single also remained protective (a OR □0.45, 95%□CI□0.22–0.92, p□=□0.028), and AZT-based regimens showed borderline significantly higher odds (aOR □3.33, 95%□CI□1.43–7.75, p□=□0.005).

**Table 4:**
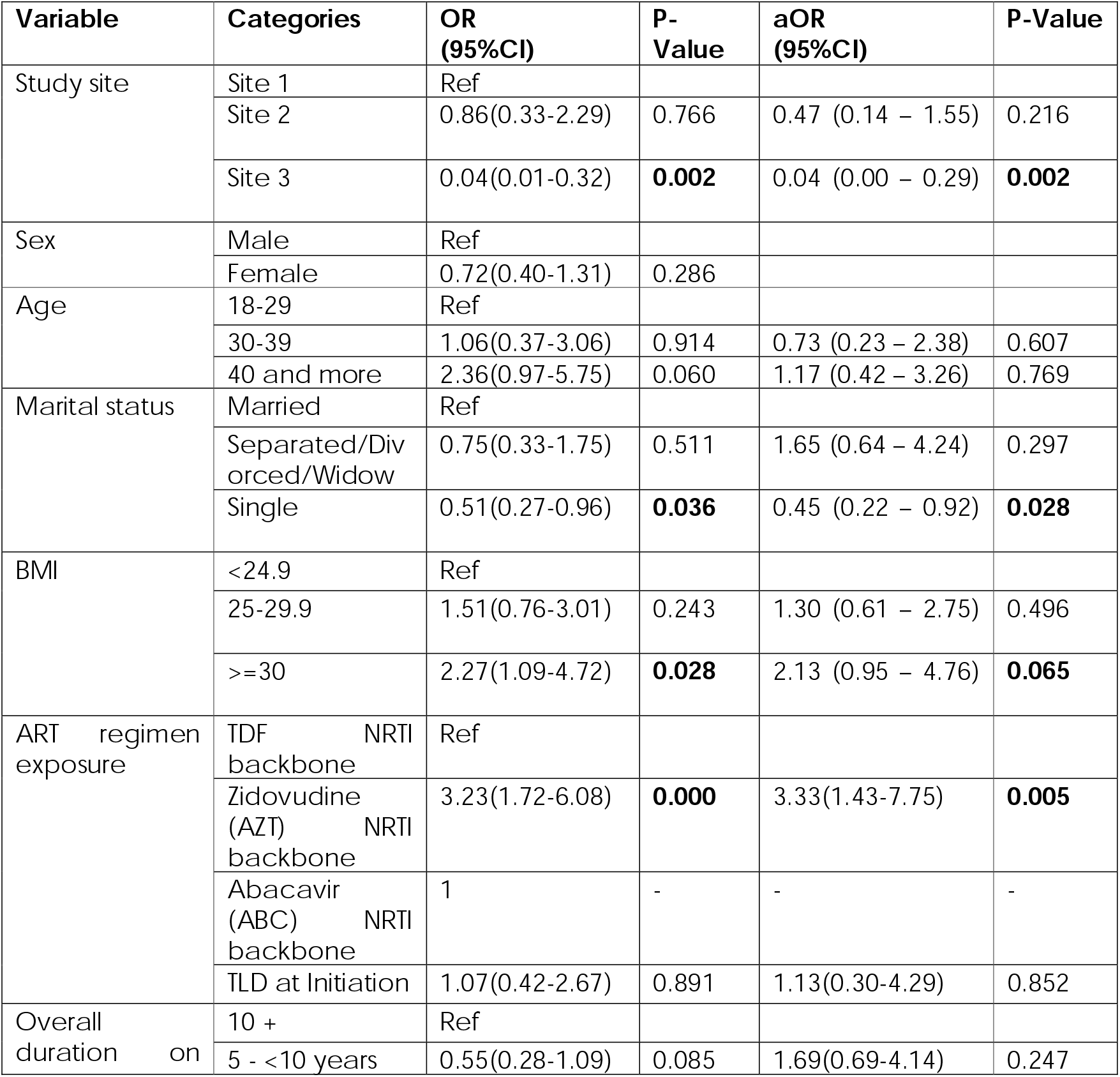
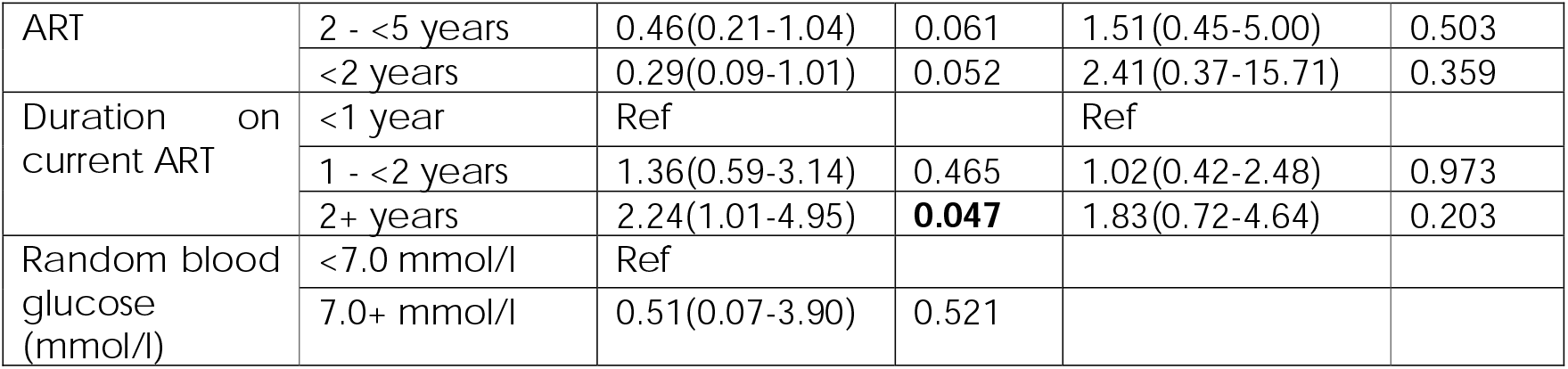
Bivariate and Multivariate Analysis for Factors Associated with adverse events.

## Discussion

This study assessed for AEs associated with DTG-based ART regimens among PLHIV in Uganda, focusing on occurrence, types, and predictors. We found an overall AE prevalence of 8.0% over six months, with neuropsychiatric disorder, cardiac disorders and Reproductive system as the most frequently reported categories. Multivariable analysis identified site variation, marital status, and prior exposure to zidovudine (AZT)-based regimens as important factors associated with AE occurrence. These findings offer important real-world insights into the safety profile of DTG-containing regimens in a sub-Saharan African context, complementing clinical trial data and informing clinical and programmatic decisions.

Our observed overall AE rate of 8.0% aligns with emerging evidence from sub ⍰Saharan Africa, where reported AE rates among patients on DTG ⍰based regimens range widely from 5% to 33% depending on study design, follow ⍰up duration, and patient profiles[15,16]. This relatively low incidence strengthens the growing consensus that DTG is generally well tolerated and safe in routine clinical settings, consistent with WHO recommendations to adopt DTG as first⍰line therapy. A plausible explanation for the low AE rate in our cohort is that most participants were ART ⍰experienced and virologically stable, which m ay have contributed to reduced susceptibility to AEs[17].

Among specific AE categories, neuropsychiatric disorders were most frequently reported (2.5%), with headaches and peripheral neuropathy predominating. This finding corroborates prior studies reporting neuropsychiatric symptoms such as insomnia, dizziness, and mood disturbances as common yet usually mild DTG ⍰related effects[15,16]. Although peripheral neuropathy has been historically linked to older agents like zidovudine or stavudine[18], its occurrence here may reflect overlapping toxicities, pre ⍰existing conditions, or comorbidities. Given that neuropsychiatric symptoms can negatively influence adherence and quality of life, systematic screening and timely management of these symptoms should be incorporated into HIV care. Cardiovascular disorders, mainly hypertension, were the second most common (1.8%). While DTG is not strongly associated with direct cardiotoxicity, hypertension remains prevalent among PLHIV[19], likely due to background metabolic risk factors, ART⍰related weight changes, or aging [6–8,20]. This finding emphasizes the importance of routine cardiovascular risk assessment and integration of prevention and treatment strategies within HIV programs after rollout of DTG based regimens[21,22].

Reproductive system and breast disorders, notably erectile dysfunction, were reported in 1.5% of participants, highlighting potential impacts on sexual health, quality of life, overall wellbeing, and adherence to therapy, particularly among men. Erectile dysfunction is multifactorial, with vascular, psychological, and medication-related contributors[23], and has been sporadically reported in association with ART. Addressing sexual health concerns is therefore essential for holistic HIV c are. Furthermore, hypogonadism is a recognized condition among HIV ⍰infected males, including those on ART, with reported prevalence ranging from 20% to 70% [24–28].These findings warrant further investigation into the underlying mechanisms and risk factors driving sexual dysfunction in this population.

Other AEs, including gastrointestinal, eye, and skin disorders, were rare (<1%), consistent with the overall favourable DTG safety profile[3]. Nonetheless, it is important to note that our monitoring primarily relied on participant awareness and reported symptomatic AEs. Incorporating objective laboratory markers such as liver enzymes, kidney function makers, lipid profiles, and glucose levels among others would enhance detection of asymptomatic but clinically significant toxicities. Conditions like hyperglycemia, dyslipidemia, or early hepatic and kidney injury may remain sub clinic al for long periods, yet when unrecognized can progress to severe complications, undermining both safety and efficacy of ART [29]. Early identification of such derangements, followed by targeted interventions, can help maintain optimal HIV control while minimizing long ⍰term morbidity.

Our analysis revealed several important predictors of AEs among participants receiving dolutegravir-based regimens. Notably, participants at site □3 (Nyimbwa Health Centre □IV) had markedly lower odds of reporting AEs compared with those at site □1 (Mildmay Uganda Hospital) (aOR □0.04). This striking difference, which persisted after adjusting for confounders, may reflect variation in catchment areas, patient demographics which may affect reporting practices. Mildmay Uganda Hospital (MUgH), as a center of excellence for HIV care and treatment, may attract more PLHIV with complex comorbidities, whereas Health Centre □IV facilities typically refer more complicated cases to higher-level centers. These lower AE reports at Health Centre IVs may therefore not necessarily indicate under-recognition but rather differences in c ase management pathways. Further investigation is needed to clarify whether enhancing AE detection and reporting practices at lower-level facilities through targeted surveillance, capacity building, or revised referral protocols to could improve comparability of AE data a cross sites..

Being single was associated with significantly reduced odds of AEs (aOR □0.45), a finding that contrasts with literature suggesting marriage often provides stronger psychosocial support and promotes adherence[30]. One possible explanation is that married individuals, often older and with longer ART exposure, may have accumulated comorbidities or treatment-related complications, increasing AE risk. Additionally, greater health ⍰seeking behavior among married participants may lead to more frequent symptom reporting compared to single counterparts. Conversely, single individuals may represent a younger, healthier subgroup with fewer underlying conditions. This unexpected pattern highlights the need for further research to explore social, behavioral, and site ⍰specific factors influencing AE reporting.

As well, participants with prior exposure to AZT-based backbones showed a strong association with AEs (aOR □3.33). AZT’s known mitochondrial and hem atologic toxicities m ay predispose patients to persistent side effects even after transitioning to TLD[18]. This highlights the importance of detailed ART history when selecting regimens and the need for closer monitoring in those with prior AZT exposure.

### Implications for Clinical Practice and Programs

Our findings highlight the overall safety and tolerability of DTG-based regimens within routine Ugandan HIV care while identifying subgroups that require closer attention. The increased AE risk among individuals with prior AZT exposure emphasizes the importance of incorporating detailed ART history into routine assessments to guide monitoring. The observed lower AE risk among single participants suggests that social and marital factors may influence symptom reporting, health-seeking behaviour, or adherence, warranting further exploration and tailored support. The notable inter ⍰site variation reinforces the need to raise awareness about AE reporting within the general population and to implement standardized detection, reporting, and management practices, ensuring consistent and uniform quality of c are across all facilities.

The predominance of neuropsychiatric, cardiovascular, and reproductive AEs emphasises the need for integrating structured symptom screening and management into routine visits. Strengthened pharmacovigilance systems with active surveillance remain essential since DTG based regimens are continually being scaled-up.

## Strengths and Limitations

This prospective cohort study employed standardized MedDRA classifications and active follow-up, enhancing the accuracy and comparability of AE data. Inclusion of sites with diverse populations from both urban and rural settings and differing ART histories provides a broad safety profile. However, potential under-reporting due to self-report bias, limited sample size for rare events, and residual confounding despite multivariable adjustment remain limitations. Selection bias occurred because participants were ART-experienced and relatively stable on therapy, which could have influenced the findings. Longer follow-up would be valuable to assess the persistence or late onset of adverse events, and including ART-naïve participants would help establish AE causality.

## Conclusion

DTG⍰based ART regimens showed a favourable safety profile among Ugandan PLHIV, with a low overall incidence of adverse events, primarily neuropsychiatric, cardiovascular, and reproductive symptoms. Martials status, prior AZT exposure and study site were associated with AE risk, emphasizing the need for individualized monitoring and strengthened awareness AE reporting, standardized pharmacovigilance across facilities. These findings support continued DTG use in line with WHO recommendations while emphasizing the importance of context⍰specific safety surveillance to optimize HIV care and treatment outcomes.

## Data Availability

All data produced in the present work are contained in the manuscript

## Acknowledgements

The authors extend their gratitude to the HIV-infected individuals, hospital administrators, staff, and healthcare workers. Their contributions at various levels were instrumental in making this study a success.

## Contributors

CA*, CA, YK, and SN2 conceived and designed the study. CA*, SN, CA, SN2, LJ, GE and JE did the data collection and interpretation. CA*, and IK participated in analysis. RM, LJ, GK, AS, SA, DNK, VN, BM and HBN participated in the interpretation of results and writing the final manuscript. All authors read and approved the final version of the manuscript.

